# Locus Coeruleus Integrity from 7T MRI Relates to Apathy and Cognition in Parkinson’s Disease and Progressive Supranuclear Palsy

**DOI:** 10.1101/2021.04.19.21255762

**Authors:** Rong Ye, Claire O’Callaghan, Catarina Rua, Frank H. Hezemans, Negin Holland, Maura Malpetti, P. Simon Jones, Roger A. Barker, Caroline H. Williams-Gray, Trevor W. Robbins, Luca Passamonti, James Rowe

## Abstract

**Objective:** The loss of noradrenergic neurons in the locus coeruleus contributes to neuropsychiatric symptoms in both Parkinson’s disease (PD) and progressive supranuclear palsy (PSP). This study aimed to characterise the spatial patterns of locus coeruleus pathological change in PD and PSP, and its relationship to cognitive and psychiatric symptoms.

**Methods:** Twenty-five patients meeting clinical criteria for idiopathic PD, fourteen probable PSP-Richardson’s syndrome and twenty-four age-matched healthy controls (HC) were recruited. All participants underwent clinical assessments for cognition and apathy, and high-resolution (0.08 mm^3^) 7T-magnetisation-transfer imaging to measure LC integrity *in vivo*. To establish spatial patterns of locus coeruleus change, we obtained sub-region summaries of mean contrast ratios and significant locus coeruleus clusters using voxelwise analyses (family-wise-corrected *p*<0.05); we further correlated the locus coeruleus contrast with measures of apathy and cognition.

**Results:** Reduced contrast was observed in the caudal locus coeruleus for both PD and PSP relative to controls, with significant regional and voxelwise differences (HC>PD, right caudal locus coeruleus, 37 voxels; HC>PSP, bilateral caudal locus coeruleus, 206 voxels). PSP and PD patients showed similar levels of locus coeruleus degeneration relative to controls, but this was spatially more extensive in PSP. Across both groups, variability of locus coeruleus integrity was associated with cognitive performance (MoCA: 200 voxels; ACE-R: 70 voxels) and apathy (Apathy Scale: 99 voxels).

**Conclusions:** The relationship between locus coeruleus structure and non-motor symptoms highlights a role for noradrenergic dysfunction across both PD and PSP, confirming the potential for noradrenergic therapeutic strategies to address transdiagnostic cognitive and behavioural features in neurodegenerative disease.

## INTRODUCTION

Parkinson’s disease (PD) and progressive supranuclear palsy (PSP) result from distinct pathologies, yet they share many motor and non-motor symptoms. These include executive dysfunction and neuropsychiatric features, with apathy^1 2^. While the causes of cognitive and neuropsychiatric symptoms in these diseases are multi-factorial, there is increasing evidence that noradrenergic dysfunction plays a prominent role^3-5^.

In both PD and PSP, the noradrenergic locus coeruleus is a site of selective vulnerability, undergoing early and severe pathological changes^6-8^. Diffuse projections enable the locus coeruleus to release noradrenaline in widespread brain regions. The effects of noradrenaline at target regions include altering the gain of neurons and promoting reorganisation of functional networks^9^. Through these actions, noradrenaline influences many cognitive functions, facilitating attentional focus and flexible shifting between goal-directed behaviours. In PD and PSP, we propose that vulnerability of locus coeruleus neurons and consequent loss of noradrenergic projections have wide-ranging impacts on cognition and behaviour^10^.

Since noradrenergic deficits are determinants of cognitive and behavioural impairments in PD and PSP, they are a promising treatment target for cognitive and neuropsychiatric symptoms. However, individual differences in brain structure and neurochemistry determine a patient’s response to noradrenergic treatment, often non-linear (U-shaped) relationships have been observed for dopaminergic and serotonergic interventions^5 11 12^. Specifically, we propose that an individual’s need for – and response to – noradrenergic treatment depends on the state of their locus-coeruleus noradrenergic system.

Progress in noradrenergic therapeutics has been hindered by limited *in vivo* characterisation of locus coeruleus degeneration. The locus coeruleus is small and elongated (∼1×16 mm) with few neurons (∼50,000 in humans), which challenges standard imaging techniques. Specialised magnetisation-transfer sequences have been developed that are sensitive to the intrinsic contrast generated by neuromelanin-rich noradrenergic cells of the locus coeruleus^13 14^. These correlate with the density of neuromelanin-positive neurons in the locus coeruleus^15^, providing a biomarker of locus coeruleus degeneration^16^ that confirms reductions in PD and PSP^17 18^. Such sequences are most sensitive at ultra-high field 7T MRI^19 20^, providing both enhanced contrast-to-noise and improved spatial resolution.

Here, we test the hypothesis that apathy and cognitive function correlate with the integrity of the locus coeruleus in PD and PSP, exploiting ultra-high field neuromelanin-sensitive 7T MRI. The high resolution (0.08 mm^3^) enabled localisation of degeneration to sub-regions of the locus coeruleus. Given the prominent role the locus coeruleus-noradrenergic system plays in cognition and goal-directed behaviours, we predicted that clinical assessments of cognition and apathy would relate to locus coeruleus integrity, with a rostro-caudal gradient of degeneration.

## METHODS

### Participants

Sixty-three participants aged between 50-80 years were recruited, including 25 people with idiopathic PD, 14 with PSP-Richardson’s syndrome and 24 age- and sex-matched healthy controls. Patients with idiopathic Parkinson’s disease were recruited via the University of Cambridge Parkinson’s Disease Research Clinic and the Parkinson’s UK volunteer network, and met the United Kingdom Parkinson’s Disease Society Brain Bank criteria for PD diagnosis. Patients with PSP were recruited from a tertiary clinic at the Cambridge University Centre for Parkinson-plus, with probable PSP-Richardson’s syndrome (Movement Disorder Society 2017 criteria). Healthy controls were recruited from local volunteer panels. No controls were using psychoactive medications and exclusion criteria for all participants included current or history of ischaemic or haemorrhagic stroke, severe co-morbidity, and any contraindications to 7T MRI. None of the patients had impulse control disorders, based on clinical impression or the Questionnaire for Impulsive-Compulsive Disorders in Parkinson’s Disease screening tool. Patients did not have dementia, based on Mini-Mental State Examination (MMSE score >26)^21^ and clinical impression of cognitive function. All PD patients and 10/14 PSP patients were taking dopaminergic medications, and Levodopa equivalent daily dose (LEDD) scores were calculated. The study was approved by the local Cambridge Research Ethics Committees and participants provided written informed consent according to the Declaration of Helsinki.

### Clinical assessments

Participants completed assessments of global cognition (MMSE, Addenbrooke’s Cognitive Examination Revised; ACE-R and Montreal Cognitive Assessment; MoCA) and self-rated mood/behaviour questionnaires: Apathy Scale^22^, Barratt Impulsiveness Scale^23^, Hamilton Anxiety and Depression Scales (noting that several questions are probing apathy not low mood), and the REM Sleep Behaviour Disorder Screening Questionnaire. Higher scores on these questionnaires indicate more severe impairment. For the patients, motor severity was assessed using the Movement Disorder Society Unified Parkinson’s Disease Rating Scale motor section (MDS-UPDRS-III). PSP patients additionally underwent the PSP Rating Scale (PSPRS). For both measures, higher scores are consistent with more severe motor symptoms. Patients underwent MRI and clinical assessments on their regular medications (see Table 1).

**Table 1.** Mean (standard deviation) for demographics and clinical assessments.

|  | Descriptive |  |  | *p* values for pairwise tests |  |  |
| --- | --- | --- | --- | --- | --- | --- |
|  | HC | PD | PSP | HC vs PD | HC vs PSP | PD vs PSP |
| **Age (years)** | 65.5 (5.5) | 67.4 (7.4) | 69.7 (7.7) | 0.579 | 0.163 | 0.576 |
| **Education (years)** | 14.8 (3.1) | 14.1 (2.3) | 12.3 (2.8) | 0.603 | 0.02 | 0.13 |
| **Male/Female** | 13/11 | 18/7 | 8/6 | 0.196 | 0.859 | 0.345 |
| **MMSE** | 29.75 (0.53) | 29.52 (0.65) | 28.5 (1.74) | 0.685 | **<0.001** | 0.007 |
| **MoCA** | 28.58 (1.44) | 27.96 (1.88) | 24 (3.94) | 0.628 | **<0.001** | **<0.001** |
| **ACER-total** | 97.71 (3.25) | 95.40 (3.61) | 87.21 (7.17) | 0.182 | **<0.001** | **<0.001** |
| **Apathy Scale** | 10.38 (5.25) | 12.44 (5.43) | 20 (9.49) | 0.508 | **<0.001** | 0.003 |
| **BIS** | 55.71 (9.56) | 58.18 (10.31) | 63.86 (12.44) | 0.692 | 0.064 | 0.248 |
| **HADS-depression** | 2.83 (2.84) | 4.24 (2.73) | 7.43 (4.27) | 0.273 | **<0.001** | 0.01 |
| **HADS-anxiety** | 4.29 (3.53) | 5.04 (3.16) | 6.57 (3.2) | 0.711 | 0.11 | 0.356 |
| **RBDSQ** | - | 5.48 (3.65) | 3.07 (1.82) | - | - | 0.027 |
| **Disease Duration (years)** | - | 5 (3.05) | 4.24 (2.68) | - | - | 0.438 |
| **LEDD** | - | 644.3 (499.36) | 323.57 (389.4) |  |  | 0.038 |
| **UPDRS-III** | - | 28.36 (11.98) | 33.07 (6.96) | - | - | 0.187 |
| **PSPRS** | - | - | 30.79 (9.11) | - | - | - |

Group difference in sex was examined using chi-square test. A one-way ANOVA was used for group difference with post-hoc Tukey HSD *p* values provided for pairwise comparisons. RBDSQ, disease duration and UPDRS-III were compared with independent samples *t*-test between PD and PSP. MMSE: Mini-Mental State Examination, MoCA: Montreal Cognitive Assessment, ACER: Addenbrooke's Cognitive Examination Revised, BIS: Barratt Impulsiveness Scale, HADS: Hamilton Anxiety and Depression Scale, RBDSQ: REM Sleep Behaviours Screening Questionnaire, LEDD: Levodopa equivalent daily dose, UPDRS: Unified Parkinson's Disease Rating Scale, PSPRS: Progressive Supranuclear Palsy Rating Scale. Significant *p* values ( $p < 0.0016$ , equivalent to $p < 0.05$ with Bonferroni correction for multiple comparisons of all tests on demographics and clinical assessments) are highlighted in bold.

### MRI acquisition

Participants underwent MR scan on a 7T Magnetom Terra (Siemens, Erlangen, Germany) with a 32-channel head coil (Nova Medical, Wilmington, USA). The locus coeruleus was imaged using a sensitive 3-D magnetisation transfer (MT) weighted sequence^19^ at high resolution (0.4 × 0.4 × 0.5 mm^3^), following the same protocol in previous studies^5 20^. A high resolution T1-weighted structural image (0.7 mm isotropic) was acquired using MP2RAGE sequence with the UK7T Network harmonised protocol^24^. For the MT-weighted sequence, 112 oblique, high-resolution, axial slices were placed perpendicular to the long axis of the brainstem, covering both midbrain and pons. A train of 20 MT pulses at 6.72 ppm off resonance were applied followed by a turbo-flash readout (TE = 4.08 ms, TR = 1251 ms, flip-angle = 8°, voxel size = 0.4 × 0.4 × 0.5 mm^3^, 6/8 phase and slice partial Fourier, bandwidth = 140 Hz/px, no acceleration, 14.3%-oversampling, TA ∼ 7 min). The MT sequence was repeated twice and averaged offline to enhance the signal-to-noise ratio. An additional scan without MT pre-saturation was acquired for registration purposes. A high resolution T1-weighted structural image was acquired using MP2RAGE sequence: TE = 2.58 ms, TR = 3500 ms, BW = 300 Hz/px, voxel size = 0.7 × 0.7 × 0.7 mm^3^, FoV = 224 × 224 × 157 mm^3^, acceleration factor (A>>P) = 3, flip angles = 5/2° and inversion times (TI) = 725/2150 ms for the first/second images.

### Locus coeruleus integrity estimation

To limit potential confounds in standard intensity-based segmentation and increase the sensitivity, an atlas-based segmentation approach was adopted to extract LC signal using a 5% probabilistic LC atlas in the standard space, generated from an independent sample of 29 older healthy volunteers. The MT images were co-registered to the high-resolution (0.5 mm isotropic) ICBM152 2009b template following a T1-driven, intra-modality coregistration pipeline^20^. FreeSurfer (v6.0.0) was used for segmenting and estimating volumes of individual brainstem substructures. Pairwise Dice Similarity Coefficients (DSC) between individual (I) coregistered and standard (S) pons were calculated with DSC=2(I∩S)/(I+S) to provide the measurement of registration accuracy. Voxelwise contrast-to-noise ratio (CNR) of the LC was computed against a 4 × 4 × 4.5 mm^3^ cubic reference region placed at the central pons (Fig1A) using the difference between a given voxel and the mean intensity of the reference region divided by the standard deviation of the reference signals. This pontine reference region was chosen as a measurement source of noise due to its proximity to the LC. Although significant pontine tau pathology has been reported in PSP-Richardson’s syndrome^25^, the MT signal did not differ between PSP patients and controls in the current study (t(36) = 0.472, *p* = 0.64), confirming this area as a suitable reference region. The independent LC atlas was applied to extract averaged CNR for each slice, three equidistantly subdivided regions (rostral, central, and caudal), left and right LC, and the whole structure. The same LC mask was also used in the voxelwise analyses.

**Figure 1.**
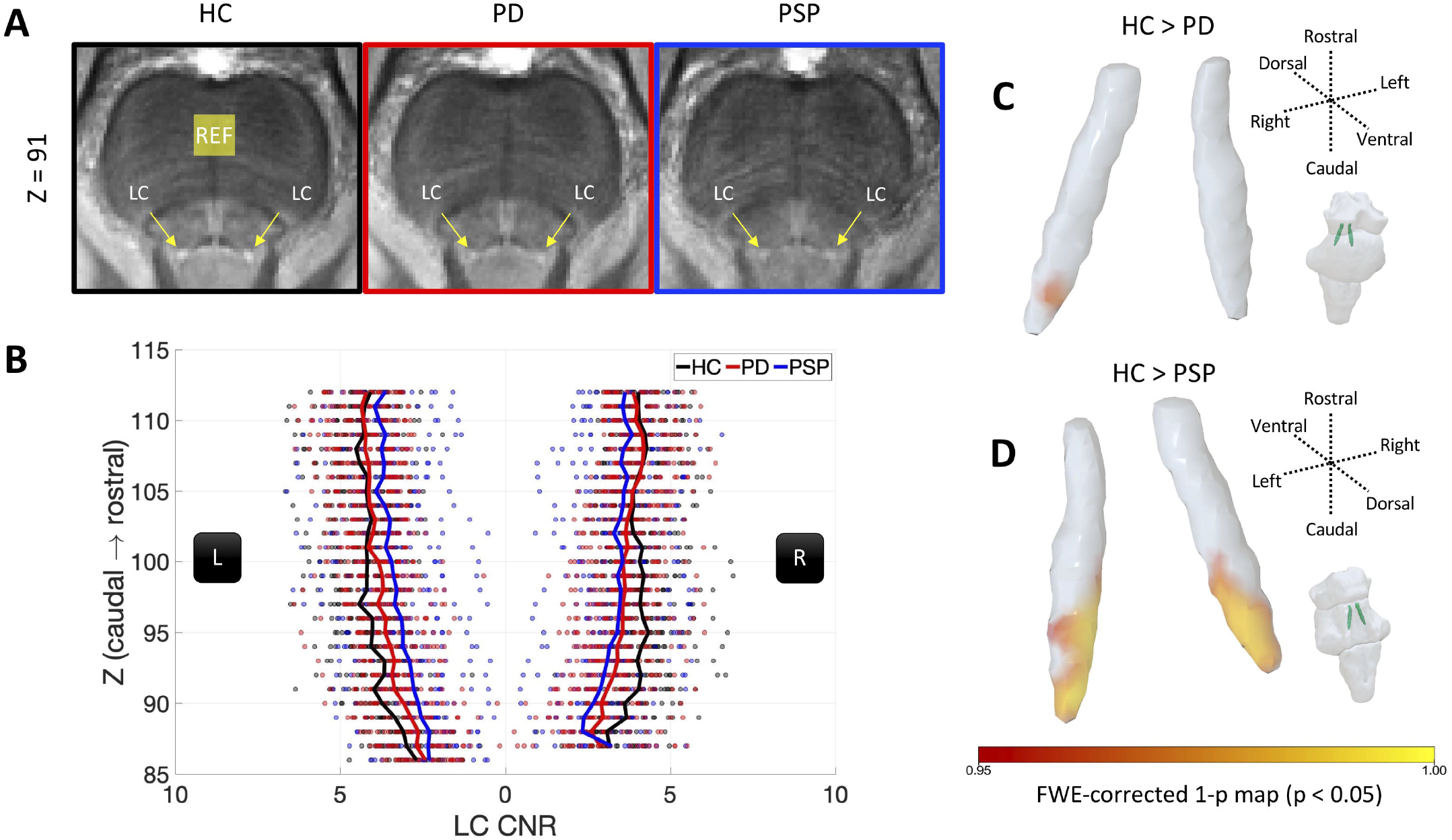
Comparisons of LC integrity across the three groups. The (A) rostrocaudal distributions of LC CNR and (B) volume (shaded area represents standard error) are presented with axial MT images (C), showing substantial contrast loss in patient groups. The voxelwise analyses further confirmed the observation where significant contrast reduction can be seen in right caudal LC for (D) PD and bilateral caudal LC for (E) PSP compared with HC (independent t-test, 10,000 permutations, FWE-corrected p<0.05).

### Voxelwise Analyses

Voxelwise analyses provide a more sensitive and spatially accurate localisation for the LC group differences and clinical correlations, as averaged CNR is insensitive to voxel-by-voxel variations in LC integrity. The co-registered CNR maps were smoothed with a 1 mm FWHM Gaussian kernel, masked with the 5% 7T LC atlas, then subjected to general linear models for t-tests of group differences. Relationships with clinical variables were assessed by regression models where clinical scores were mean-centred across patient groups. Voxelwise analyses used the *randomise* function in FSL (v6.0.0). A threshold free cluster enhancement (TFCE) method was adopted for cluster inference combined with permutation tests (10,000 iterations). Analogous regression models included age and LEDD as covariates of no interest to test potential confounds because age is a significant contributor to the pathogenesis and cognitive performance might be affected by dopaminergic medication. The family-wise error (FWE) corrected p-value (<0.05) was used to determine significant clusters.

### Statistical analyses of LC integrity

Statistical analyses were performed in JASP (v0.11.1) and R (version 3.6.1). Group differences in clinical assessments and LC contrast across the subregions were tested using ANOVAs. The relationships between LC contrast and clinical assessments were validated using regional mean LC CNR in patient groups with linear regression models with clinical scores as the dependent variable and LC CNR as a fixed effect. We report the results for the hypothesis testing with a significance level of *p* = 0.05.

## RESULTS

### Demographics and Clinical Assessment

Demographics and clinical data are summarised in Table 1. Controls were age- and sex-matched to patient groups. Patients with PSP had fewer years of education compared to controls, and they were more impaired on the MMSE, MoCA, ACE-R, Apathy Scale and HADS-depression assessments relative to both controls and PD. Patients with PD had higher LEDD and RBDSQ scores than PSP.

### Locus Coeruleus Group Differences

LC integrity assessment benefited from the atlas-based approach, which has clear advantages over classic segmentation-based methods: pathology-related LC signal reduction is accompanied by increased segmentation failure and reduced spatial precision, leading to inflated contrast estimation. Using regional averaged LC contrast, there were group differences in the caudal subregion (F(2,60) = 4.05, *p* = 0.02) between PSP patients and controls (*p*_*tukey*_ = 0.02), but not for PD versus controls (*p*_*tukey*_ = 0.16) or PD versus PSP (*p*_*tukey*_ = 0.48). There was no difference across the three groups in whole-LC contrast (F(2,60) = 1.759, *p* = 0.181) or other subregions (rostral: F(2,60) = 1.066, *p* = 0.351; central: F(2,60) = 1.293, *p* = 0.282).

There was no significant effect of lateralisation on whole-LC contrast (F(1,60) = 3.802, *p* = 0.06), or group × lateralisation interaction (F(2,60) = 2.637, *p* = 0.08), although in view of the weak trend we tested and confirmed a marginal left-lateralised effect of LC contrast in PD (left > right: *p*_*holm*_ = 0.05). To examine the correlation between the asymmetries of LC contrast and motor symptoms, we created two variables for PD patients categorising the side (left or right) of lateralisation for LC atrophy (lower contrast) in each subregion and more affected side (left or right) for motor symptoms by weighing scores for the left and right side from the MDS-UPDRS-III (higher scores for more severe symptoms). Chi-square tests were used to test the independency and Cramer’s V was used to calculate the correlation coefficient for two categorical variables. Our results showed that lower contrast on one side of rostral and central LC was consistent with more severe motor symptoms ipsilaterally (rostral: χ^2^ = 5.24, *p* = 0.02, Cramer’s V = 0.46; central: χ^2^ = 3.74, *p* = 0.05, Cramer’s V = 0.39; caudal: χ^2^ = 0.43, *p* = 0.51, Cramer’s V = 0.13:). We also calculated two asymmetry indexes (AI) for motor symptoms and whole-LC contrast, respectively. AI was defined as 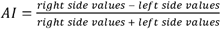 There was a positive correlation between motor-AI and LC-AI in PD patients (R^2^ = 0.22 p = 0.019), suggesting a relationship between lateralisation of LC contrast and motor symptoms.

The whole-LC contrast did not differ by sex (t(62) = 0.7, *p* = 0.48). There was no linear nor quadratic relationship detected between LC contrast and age (linear: F(1,61) = 0.84, R^2^ = 0.01, *p* = 0.36; quadratic: F(2,60) = 1.55, R^2^ = 0.05, *p* = 0.16).

To ensure that the caudal findings were not driven by varied noise levels in subregions or across groups, we compared the signal-to-noise ratio (SNR) for three subregions and examined the subregion × group interaction. The SNR differed between subregions (F(2,120) = 13.02, *p* < 0.001). However, the effect was only evident in the rostral LC (LC_rostral_ > LC_central_: *p*_*holm*_ < 0.001; LC_rostral_ > LC_caudal_: *p*_*holm*_ = 0.003), whereas SNRs of central and caudal LC did not differ (*p*_*holm*_ = 0.525). There was no group by subregion SNR interaction (F(4,120) = 2.22, *p* = 0.07). Furthermore, there was no group difference observed for pons volumes (F(2,59) = 2.27, *p* = 0.11) or LC registration accuracy estimated using Dice Similarity Score (DSC) on individual pons masks segmented from coregistered T1 images (DSC Mean ± SD: 0.875 ± 0.02, range: 0.74 - 0.89, F(2,60) = 1.388, *p* = 0.258), indicating that the contrast difference in the LC was not attributed to these potential confounds. Adding both caudal LC SNR and registration accuracy did not meaningfully change the caudal findings in patients (F(2,56) = 4.63, *p* = 0.01; HC vs PSP: *p*_*tukey*_ = 0.01; HC vs PD: *p*_*tukey*_ = 0.38; PD vs PSP: *p*_*tukey*_ = 0.2).

More sensitive voxelwise analyses revealed clusters with significantly reduced contrast (threshold free cluster enhancement, *p*<0.05, FWE-corrected) in both disease groups compared to controls (Table 2). Focal LC atrophy was identified for PD patients in the right caudal LC (Fig 1C, 37 voxels) whereas PSP patients had extensive bilateral caudal damage in the LC (Fig 1D, 206 voxels). There was no group difference between PD and PSP by voxelwise analysis.

**Table 2.** TFCE clusters within the LC for regression models and group comparisons (FWE-corrected p<0.05)

|  | coordinates of local maxima (mm) |  |  |  | volume |  |
| --- | --- | --- | --- | --- | --- | --- |
|  | x | y | z | voxel | (mm<sup>3</sup>) | t |
| Regression |  |  |  |  |  |  |
| MoCA | -3 | -38.5 | -22.5 | 116 | 14.5 | 2.9 |
|  | 4.5 | -39.5 | -26.5 | 84 | 10.5 | 2.95 |
| ACE-R | -4 | -39.5 | -26 | 63 | 7.875 | 3.01 |
|  | 4.5 | -39.5 | -26.5 | 3 | 0.375 | 3.23 |
|  | 3.5 | -39 | -23.5 | 3 | 0.375 | 3.12 |
|  | 2 | -37.5 | -19 | 1 | 0.125 | 3.2 |
| AS | -4.5 | -39 | -22.5 | 99 | 12.375 | 2.54 |
| Regression with<br>Age and LEDD<br>covariates |  |  |  |  |  |  |
| MoCA | 4.5 | -39.5 | -26 | 107 | 13.375 | 4.42 |
|  | -3 | -38.5 | -22.5 | 103 | 12.875 | 4.51 |
|  | -3.5 | -36 | -16.5 | 2 | 0.25 | 3.51 |
| ACE-R | -3 | -38.5 | -22.5 | 57 | 7.125 | 4.66 |
|  | 2 | -37.5 | -19 | 35 | 4.375 | 3.44 |
| AS | -2.5 | -37.5 | -18 | 55 | 6.875 | 3.32 |
|  | -5 | -39.5 | -26 | 21 | 2.625 | 2.92 |
| HC > PSP | -5 | -40 | -27.5 | 124 | 15.5 | 2.9 |
|  | 3.5 | -39 | -25.5 | 82 | 10.25 | 2.92 |
| HC > PD | 4.5 | -38.5 | -27 | 37 | 4.625 | 2.93 |

### Locus Coeruleus Integrity and Clinical Assessments in Patients

Using averaged summary scores of LC contrast, MoCA performance was associated with whole LC contrast (Fig 2A, F(1,37) = 5.34, R^2^ = 0.13, *p* = 0.03), and the ACE-R and Apathy scores were associated with left LC contrast values (ACE-R: Fig 2B, F(1,37) = 4.3, R^2^ = 0.1, *p* = 0.04; Apathy Scale: Fig 2C, F(1,37) = 4.34, R^2^ = 0.08, *p* = 0.04) in patient groups. A significant group × LC interaction was detected only for Apathy scores (*p* = 0.01), where the effect was present in PSP (F(1,12) = 5.75, *p* = 0.03) but not significant for the PD group (F(1,23) = 0.93, *p* = 0.34). There was no group effect observed for the relationships between LC integrity and MoCA (*p* = 0.93) or ACE-R (*p* = 0.67) assessments.

**Figure 2.**
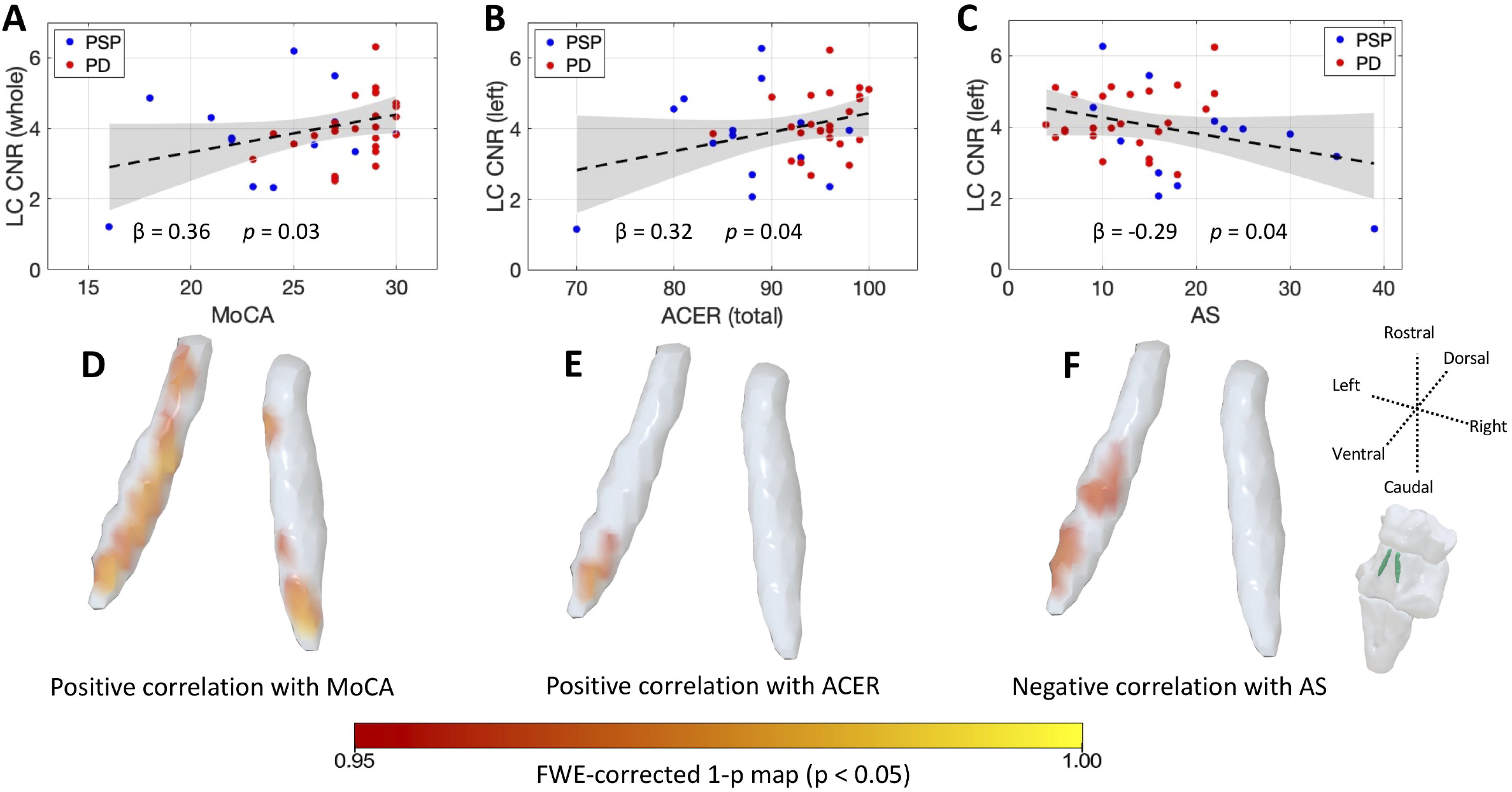
The role of LC in global cognition and apathy. The MoCA (A), ACE-R (B) and AS Apathy score (C) scores in PD and PSP correlate with LC contrast. The spatial distributions within the LC contributing to global cognition and apathy are revealed from voxelwise regression models (D-F, 10,000 permutations, FWE-corrected p<0.05).

Across all patients, the LC contrast was not associated with UPDRS motor scores (F(1,37) = 0.09, R^2^ < 0.01, *p* = 0.77), disease duration (F(1,37) = 0.02, R^2^ < 0.01, *p* = 0.9) or RBDSQ (F(1,37) = 0.77, R^2^ = 0.02, *p* = 0.39). Marginally significant relationships were observed between the LC contrast and BIS scores (F(1,37) = 3.87, R^2^ = 0.08, *p* = 0.06), Hamilton anxiety (F(1,37) = 4.27, R^2^ = 0.1, *p* = 0.05) and depression scores (F(1,37) = 4.03, R^2^ = 0.1, *p* = 0.05). For cases with PSP, the LC contrast was not correlated with the PSPRS scores (F(1,12) = 1.5, R^2^ = 0.11, *p* = 0.24).

A voxelwise analysis would have advantages for detecting and localising the effect of LC in cognition and apathy if there are localised effects or if the arbitrary division of the LC into three parts did not correspond to neuropathological divisions. The voxel-wise results confirmed bilateral LC clusters were positively correlated with MoCA scores (Fig 2D; TFCE, FWE-corrected *p*<0.05), and higher contrast in the left caudal LC clusters was positively correlated with ACE-R scores (Fig 2E): these positive correlations indicate preserved LC integrity was associated with better cognitive performance. Consistently, the left central and caudal LC clusters were negatively correlated with Apathy (Fig 2F), thus reduced LC integrity was associated with worse apathy. Adding age and LEDD as covariates of no interest did not meaningfully change the results of voxelwise analyses (Table 2).

## DISCUSSION

Using ultra-high field 7T magnetisation-transfer imaging, we confirm *in vivo* loss of locus coeruleus integrity in Parkinson’s disease and Progressive Supranuclear Palsy, both disorders with previously well-established locus coeruleus pathology at *post mortem*. We confirm the regionally specific pattern of predominantly caudal locus coeruleus changes in PD and PSP, which was associated with cognitive impairment and apathy. These findings highlight the importance of the locus coeruleus-noradrenaline system in cognitive and behavioural features of these diseases. They demonstrate the utility of neuromelanin-sensitive imaging at ultra-high field to measure the locus coeruleus in neurodegenerative disease, which we suggest will be an important biomarker for use in future clinical trials, where patients could be stratified according to locus coeruleus integrity^5 20^.

The main group-wise effect of PD and PSP on the locus coeruleus was most apparent in the caudal subregion. This rostro-caudal gradient has been noted in some neuropathological studies of PD^26^, although not all^27^, and is in keeping with more recent *in vivo* LC imaging showing a prominence of caudal changes in PD^28 29^. In contrast, Alzheimer’s disease, Down’s syndrome^27^, and healthy ageing^30^ have been associated with greater cell loss from the rostral locus coeruleus. The caudal sub-region may be especially vulnerable as it lies adjacent to the 4^th^ ventricle, exposed to environmental insults from the cerebrospinal fluid^31^. Also, the caudal locus coeruleus receives projections from the vagal nerve via the nucleus of the solitary tract which, speculatively, make it more vulnerable to transmission of misfolded proteins from the periphery^32^. Future studies would be required to establish the relationship to peripheral molecular pathology.

In our PD and PSP participants, the relationship of LC signal to behavioural (Apathy) and cognitive (MoCA) change was not confined to caudal regions, but extended to mid and rostral LC (Fig 2). Locus coeruleus degeneration and reduced noradrenaline in neurodegenerative disease is linked with a higher risk of progression to dementia^4 33^, and faster cognitive decline in older adults^34^. The correlation of apathy to LC integrity – most clearly in the PSP group – is in keeping with extensive human and preclinical evidence linking goal directed behaviour with the noradrenergic system^35 36^. Our results highlight that whilst cognitive and motivation deficits in neurodegenerative disease have multi-factorial causes, the locus coeruleus-noradrenaline system plays a key role via its widespread modulation of target structures supporting these processes.

Cognitive decline and disorders of goal directed behaviour, including the co-occurrence of both apathy and impulsivity^37 38^, are common in both PD and PSP. The link we have shown between locus coeruleus integrity and these non-motor symptoms supports the rationale for noradrenergic therapies in selected patients. Preliminary work in PD suggests that the noradrenergic reuptake inhibitor atomoxetine can improve goal directed behaviour in some people^12^, where drug responsivity depends on locus coeruleus integrity^5^. The ability to detect localised changes in the locus coeruleus as demonstrated here in PD and PSP, suggests the possibility to stratify patients in future clinical trials of noradrenergic therapy of cognitive and behavioural symptoms of neurodegenerative disease.

The role of LC in motor control is less clear. We did not find an overall significant association between LC integrity and motor symptom severity in PD, although reduced signal in the rostral and central LC portions was associated with more severe motor symptoms ipsilaterally. This is consistent with recent evidence of lateralisation of LC changes to the side of the body more affected by motor symptoms^29^. Given LC ascending projections to ipsilateral motor regions^39^, LC changes may affect motor pathways on the clinically-defined most affected side. However, the association may be indirect, with ipsilateral propagation of alpha-synuclein pathology leading to correlated pathology in both LC and motor systems, without a direct causal mechanism^29 40^.

The current study exploited the benefit of ultra-high field 7T MRI, for assessment of the locus coeruleus *in vivo*. The short imaging sequence^19^ was well-tolerated by people with PD and PSP. For locus coeruleus signal extraction, we advanced a cross-modality co-registration pipeline and an atlas-based approach, to maximise spatial precision and avoid estimation biases that occur with classic segmentation-based methods. Potential confounds specific to locus coeruleus imaging were also controlled and explicitly tested, including the consistency of noise level along the rostrocaudal axis and the effect of age. Moreover, the spatial localisation of effects was further improved by voxelwise analyses that exploited the high resolution afforded by 7T MRI. Not only do these advances allow for more accurate locus coeruleus quantification, they permit the localisation of changes to locus coeruleus sub-regions, which have previously been identified in post mortem studies^27^. The histological validation of neuromelanin-sensitive imaging^15^ provides additional evidence that the results we report here reflect the underlying pathological changes that have been observed at *post mortem*. Together, these findings pave the way for future clinical studies to investigate contributions of the locus coeruleus-noradrenaline system in the pathogenesis, progression and treatment of neurodegenerative diseases.

## CONCLUSION

The locus coeruleus is a site of selective vulnerability to neurodegenerative disease^10 16^. We provide *in vivo* confirmation of locus coeruleus degeneration in Parkinson’s disease and Progressive Supranuclear Palsy and demonstrate its association with cognition and apathy. Ultra-high field 7T neuromelanin-sensitive imaging provides a valuable tool for investigating non-motor symptoms and has potential for stratifying patients in clinical trials of noradrenergic therapy.

## Data Availability

The MT and MP2RAGE data used in this study are available for non-commercial academic purposes upon request.

## Acknowledgements

This study was supported by Parkinson’s UK (K-1702); the Cambridge Centre for Parkinson-Plus; the China Scholarship Council; a Neil Hamilton Fairley Fellowship from the Australian National Health and Medical Research Council (GNT1091310); a Cambridge Trust Vice-Chancellor’s Award and Fitzwilliam College Scholarship; the Association of British Neurologists – Patrick Berthoud Charitable Trust (RG99368); the Medical Research Council (SUAG/051 G101400; MR/P01271X/1); James S. McDonnell Foundation 21st Century Science Initiative Scholar Award in Understanding Human Cognition; the Wellcome Trust (103838); a RCUK/UKRI Research Innovation Fellowship awarded by the Medical Research Council (MR/R007446/1); the NIHR Cambridge Clinical Research Facility and the NIHR Cambridge Biomedical Research Centre (BRC-1215-20014). For the purpose of open access, the author has applied a CC BY public copyright licence to any Author Accepted Manuscript version arising from this submission. The views expressed are those of the authors and not necessarily those of the NHS, the NIHR or the Department of Health and Social Care.

## Author Contributions

R.Y., C.O., T.W.R., L.P. and J.B.R. contributed to the conception and design of the study. M.M., C.O., C.R., F.H.H., P.S.J., N.H., M.M., C.T.R., R.A.B. and C.H.W.G. contributed to the acquisition and analysis of data. R.Y., C.O., L.P. and J.B.R. contributed to drafting the text and preparing the figures.

## REFERENCES

1. Kehagia AA, Barker RA, Robbins TW. Neuropsychological and clinical heterogeneity of cognitive impairment and dementia in patients with Parkinson’s disease. Lancet Neurol 2010;9(12):1200–13. doi: 10.1016/S1474-4422(10)70212-X [published Online First: 2010/10/01]

2. Aarsland D, Litvan I, Larsen JP. Neuropsychiatric symptoms of patients with progressive supranuclear palsy and Parkinson’s disease. J Neuropsychiatry Clin Neurosci 2001;13(1):42–9. doi: 10.1176/jnp.13.1.42 [published Online First: 2001/02/24]

3. Passamonti L, Lansdall CJ, Rowe JB. The neuroanatomical and neurochemical basis of apathy and impulsivity in frontotemporal lobar degeneration. Curr Opin Behav Sci 2018;22:14–20. doi: 10.1016/j.cobeha.2017.12.015 [published Online First: 2019/04/30]

4. Halliday GM, Leverenz JB, Schneider JS, et al. The neurobiological basis of cognitive impairment in Parkinson’s disease. Mov Disord 2014;29(5):634–50. doi: 10.1002/mds.25857 [published Online First: 2014/04/24]

5. O’Callaghan C, Hezemans FH, Ye R, et al. Locus coeruleus integrity and the effect of atomoxetine on response inhibition in Parkinson’s disease. Brain 2021;144(8):2513–26. doi: 10.1093/brain/awab142 [published Online First: 2021/03/31]

6. Braak H, Del Tredici K, Rub U, et al. Staging of brain pathology related to sporadic Parkinson’s disease. Neurobiol Aging 2003;24(2):197–211. doi: 10.1016/s0197-4580(02)00065-9 [published Online First: 2002/12/25]

7. Surmeier DJ, Obeso JA, Halliday GM. Selective neuronal vulnerability in Parkinson disease. Nat Rev Neurosci 2017;18(2):101–13. doi: 10.1038/nrn.2016.178 [published Online First: 2017/01/21]

8. Kaalund SS, Passamonti L, Allinson KSJ, et al. Locus coeruleus pathology in progressive supranuclear palsy, and its relation to disease severity. Acta Neuropathol Commun 2020;8(1):11. doi: 10.1186/s40478-020-0886-0 [published Online First: 2020/02/06]

9. Aston-Jones G, Cohen JD. An integrative theory of locus coeruleus-norepinephrine function: adaptive gain and optimal performance. Annu Rev Neurosci 2005;28:403–50. doi: 10.1146/annurev.neuro.28.061604.135709 [published Online First: 2005/07/19]

10. Holland N, Robbins TW, Rowe JB. The role of noradrenaline in cognition and cognitive disorders. Brain 2021 doi: 10.1093/brain/awab111

11. Ye Z, Altena E, Nombela C, et al. Selective serotonin reuptake inhibition modulates response inhibition in Parkinson’s disease. Brain 2014;137(Pt 4):1145–55. doi: 10.1093/brain/awu032 [published Online First: 2014/03/01]

12. Ye Z, Altena E, Nombela C, et al. Improving response inhibition in Parkinson’s disease with atomoxetine. Biol Psychiatry 2015;77(8):740–8. doi: 10.1016/j.biopsych.2014.01.024 [published Online First: 2014/03/25]

13. Keren NI, Lozar CT, Harris KC, et al. In vivo mapping of the human locus coeruleus. Neuroimage 2009;47(4):1261–7. doi: 10.1016/j.neuroimage.2009.06.012 [published Online First: 2009/06/16]

14. Betts MJ, Cardenas-Blanco A, Kanowski M, et al. In vivo MRI assessment of the human locus coeruleus along its rostrocaudal extent in young and older adults. Neuroimage 2017;163:150–59. doi: 10.1016/j.neuroimage.2017.09.042 [published Online First: 2017/09/26]

15. Keren NI, Taheri S, Vazey EM, et al. Histologic validation of locus coeruleus MRI contrast in post-mortem tissue. Neuroimage 2015;113:235–45. doi: 10.1016/j.neuroimage.2015.03.020 [published Online First: 2015/03/21]

16. Betts MJ, Kirilina E, Otaduy MCG, et al. Locus coeruleus imaging as a biomarker for noradrenergic dysfunction in neurodegenerative diseases. Brain 2019;142(9):2558–71. doi: 10.1093/brain/awz193 [published Online First: 2019/07/22]

17. Ohtsuka C, Sasaki M, Konno K, et al. Differentiation of early-stage parkinsonisms using neuromelanin-sensitive magnetic resonance imaging. Parkinsonism Relat Disord 2014;20(7):755–60. doi: 10.1016/j.parkreldis.2014.04.005 [published Online First: 2014/04/29]

18. Wang J, Li Y, Huang Z, et al. Neuromelanin-sensitive magnetic resonance imaging features of the substantia nigra and locus coeruleus in de novo Parkinson’s disease and its phenotypes. Eur J Neurol 2018;25(7):949–e73. doi: 10.1111/ene.13628 [published Online First: 2018/03/10]

19. Priovoulos N, Jacobs HIL, Ivanov D, et al. High-resolution in vivo imaging of human locus coeruleus by magnetization transfer MRI at 3T and 7T. Neuroimage 2018;168:427–36. doi: 10.1016/j.neuroimage.2017.07.045 [published Online First: 2017/07/27]

20. Ye R, Rua C, O’Callaghan C, et al. An in vivo probabilistic atlas of the human locus coeruleus at ultra-high field. Neuroimage 2021;225:117487. doi: 10.1016/j.neuroimage.2020.117487 [published Online First: 2020/11/10]

21. Martinez-Martin P, Falup-Pecurariu C, Rodriguez-Blazquez C, et al. Dementia associated with Parkinson’s disease: applying the Movement Disorder Society Task Force criteria. Parkinsonism Relat Disord 2011;17(8):621–4. doi: 10.1016/j.parkreldis.2011.05.017 [published Online First: 2011/06/21]

22. Starkstein SE, Mayberg HS, Preziosi TJ, et al. Reliability, validity, and clinical correlates of apathy in Parkinson’s disease. J Neuropsychiatry Clin Neurosci 1992;4(2):134–9. doi: 10.1176/jnp.4.2.134 [published Online First: 1992/01/01]

23. Patton JH, Stanford MS, Barratt ES. Factor structure of the Barratt impulsiveness scale. J Clin Psychol 1995;51(6):768–74. doi: 10.1002/1097-4679(199511)51:6<768::aid-jclp2270510607>3.0.co;2-1 [published Online First: 1995/11/01]

24. Clarke WT, Mougin O, Driver ID, et al. Multi-site harmonization of 7 tesla MRI neuroimaging protocols. Neuroimage 2020;206:116335. doi: 10.1016/j.neuroimage.2019.116335 [published Online First: 2019/11/13]

25. Williams DR, Lees AJ. Progressive supranuclear palsy: clinicopathological concepts and diagnostic challenges. Lancet Neurol 2009;8(3):270–9. doi: 10.1016/S1474-4422(09)70042-0 [published Online First: 2009/02/24]

26. Bertrand E, Lechowicz W, Szpak GM, et al. Qualitative and quantitative analysis of locus coeruleus neurons in Parkinson’s disease. Folia Neuropathol 1997;35(2):80–6. [published Online First: 1997/01/01]

27. German DC, Manaye KF, White CL, 3rd, et al. Disease-specific patterns of locus coeruleus cell loss. Ann Neurol 1992;32(5):667–76. doi: 10.1002/ana.410320510 [published Online First: 1992/11/01]

28. Madelung CF, Meder D, Fuglsang SA, et al. The locus coeruleus shows a spatial pattern of structural disintegration in Parkinson’s disease. 2021

29. Doppler CEJ, Kinnerup MB, Brune C, et al. Regional locus coeruleus degeneration is uncoupled from noradrenergic terminal loss in Parkinson’s disease. Brain 2021 doi: 10.1093/brain/awab236 [published Online First: 2021/07/02]

30. Manaye KF, McIntire DD, Mann DM, et al. Locus coeruleus cell loss in the aging human brain: a non-random process. J Comp Neurol 1995;358(1):79–87. doi: 10.1002/cne.903580105 [published Online First: 1995/07/17]

31. Matchett BJ, Grinberg LT, Theofilas P, et al. The mechanistic link between selective vulnerability of the locus coeruleus and neurodegeneration in Alzheimer’s disease. Acta Neuropathol 2021 doi: 10.1007/s00401-020-02248-1 [published Online First: 2021/01/12]

32. Wood H. New models show gut-brain transmission of Parkinson disease pathology. Nat Rev Neurol 2019;15(9):491. doi: 10.1038/s41582-019-0241-x [published Online First: 2019/07/18]

33. Weinshenker D. Long Road to Ruin: Noradrenergic Dysfunction in Neurodegenerative Disease. Trends Neurosci 2018;41(4):211–23. doi: 10.1016/j.tins.2018.01.010 [published Online First: 2018/02/25]

34. Jacobs HI, Wiese S, van de Ven V, et al. Relevance of parahippocampal-locus coeruleus connectivity to memory in early dementia. Neurobiol Aging 2015;36(2):618–26. doi: 10.1016/j.neurobiolaging.2014.10.041 [published Online First: 2014/12/01]

35. Varazzani C, San-Galli A, Gilardeau S, et al. Noradrenaline and dopamine neurons in the reward/effort trade-off: a direct electrophysiological comparison in behaving monkeys. J Neurosci 2015;35(20):7866–77. doi: 10.1523/JNEUROSCI.0454-15.2015 [published Online First: 2015/05/23]

36. Bouret S, Richmond BJ. Sensitivity of locus ceruleus neurons to reward value for goal-directed actions. J Neurosci 2015;35(9):4005–14. doi: 10.1523/JNEUROSCI.4553-14.2015 [published Online First: 2015/03/06]

37. Scott BM, Eisinger RS, Burns MR, et al. Co-occurrence of apathy and impulse control disorders in Parkinson disease. Neurology 2020;95(20):e2769–e80. doi: 10.1212/WNL.0000000000010965 [published Online First: 2020/10/03]

38. Lansdall CJ, Coyle-Gilchrist ITS, Jones PS, et al. Apathy and impulsivity in frontotemporal lobar degeneration syndromes. Brain 2017;140(6):1792–807. doi: 10.1093/brain/awx101 [published Online First: 2017/05/10]

39. Jones BE, Moore RY. Ascending projections of the locus coeruleus in the rat. II. Autoradiographic study. Brain Res 1977;127(1):25–53. [published Online First: 1977/05/20]

40. Borghammer P. The alpha-Synuclein Origin and Connectome Model (SOC Model) of Parkinson’s Disease: Explaining Motor Asymmetry, Non-Motor Phenotypes, and Cognitive Decline. J Parkinsons Dis 2021;11(2):455–74. doi: 10.3233/JPD-202481 [published Online First: 2021/w03/09]

